# Stability of spectral results in cardiac dual-source photon-counting CT

**DOI:** 10.1101/2022.12.07.22283222

**Authors:** Leening P. Liu, Nadav Shapira, Pooyan Sahbaee, Grace J. Gang, Friedrich D. Knollman, Marcus Y. Chen, Harold I. Litt, Peter B. Noël

**Affiliations:** Department of Radiology, University of Pennsylvania, Philadelphia, PA, USA; Department of Bioengineering, University of Pennsylvania, Philadelphia, PA, USA; Siemens Healthineers, Malvern, PA, USA; National Heart, Lung, and Blood Institute, Bethesda, MD, USA; Department of Diagnostic and Interventional Radiology, School of Medicine & Klinikum rechts der Isar, Technical University of Munich, 81675 München, Germany

**Keywords:** CT, cardiac CT, photon-counting CT, spectral CT

## Abstract

**Objective:** Evaluate stability of spectral results at different heart rates, acquisition modes, and cardiac phases in first-generation clinical dual-source photon-counting CT (PCCT).

**Materials and Methods:** A cardiac motion simulator with a coronary stenosis mimicking a 50% eccentric calcium plaque was scanned with a first-generation dual-source PCCT at five different heart rates (0, 60, 70, 80, 100 bpm). Scans were performed at 120 kVp with the three available cardiac scan modes (high pitch prospectively ECG-triggered spiral, prospectively ECG-triggered axial, retrospectively ECG-gated spiral). Subsequently, virtual monoenergetic images at 50, 70, and 150 keV and iodine density maps were reconstructed at both diastole and systole to investigate the effect of the cardiac phase. Full width half max (FWHM) of the stenosis, Dice score (DSC) for the stenosed region, and eccentricity of the non-stenosed region were analyzed.

**Results:** FWHM exhibited average differences from the static FWHM across cardiac phase and heart rates of -0.20, -0.28, and -0.15 mm at VMI 150 keV for high pitch prospectively ECG-triggered spiral, prospectively ECG-triggered axial, and retrospectively ECG-gated spiral scans, respectively. DSC demonstrated similarity among parameters with standard deviations of 0.08, 0.09, 0.11, and 0.08 for VMI 50, 70, and 150 keV, and iodine density maps, respectively, with larger differences present at systole and with high pitch scans. Similarly, eccentricity illustrated small differences across heart rate and acquisition mode for each spectral result.

**Conclusions:** Consistency of spectral results at different heart rates and acquisition modes for different cardiac phase demonstrates the added benefit of spectral results from PCCT to dual-source CT to further increase confidence in quantification and advance cardiovascular diagnostics.

## Introduction

ECG-synchronized computed tomography angiography (CTA) has become an increasingly vital diagnostic tool for cardiovascular disease^1^. CTA is utilized to detect abnormal structures such as valve vegetations/calcifications, aortic dissection, and aneurysms^2–4^. It can also be used to determine the risk of heart disease and to identify coronary stenoses^5–7^. Technological advances have increased its utility and resulted in a shift in clinical day-to-day routine^8–10^. For example, for patients with stable chest pain, coronary CT angiography has shown excellent accuracy compared to catheter-based coronary angiography without the invasive nature^11^. In addition, development of different acquisition modes^12,13^ and technological advancements, such as dual source CT^14^, have addressed radiation dose exposure and temporal resolution concerns associated with retrospective low-pitch helical (spiral) acquisitions.

In dual source CT, an additional x-ray source and detector set is located approximately 90 degrees from the first x-ray source and detector. As a result of this addition, the temporal resolution is effectively doubled since the rotation required to acquire the needed data is reduced by half. This improved temporal resolution has resulted in better image quality for existing acquisition modes, especially at higher heart rates where increased motion increases the likelihood of motion-related artifacts^15^, but also enables a new acquisition mode, prospective high pitch helical (flash), which takes advantage of the two sources^16^. This mode can significantly reduce radiation dose in comparison to spiral and prospective step-and-shoot (sequence) but limits retrospective reconstruction capabilities and is most effective at low heart rates^16,17^.

Spectral CT provides additional quantitative data in cardiac CT, including material maps, that are valuable for diagnostic assessments. Photon counting CT (PCCT), the most recent realization of spectral CT, makes quantitative maps available for cardiac imaging with reduced noise, stability at low doses, increased contrast to noise ratio, and high spatial resolution in prototype and clinical PCCT scanners compared to dual energy CT^2,18–22^. Iodine density maps separate the iodine signal for the visualization of perfusion and vascular structures^23–25^ and can also be analyzed quantitatively to characterize lesions, i.e. cysts vs. hepatocellular carcinoma^26,27^. Additionally, virtual non-contrast images can obviate the need for an additional true non-contrast scan^28,29^. Virtual monoenergetic images (VMI) can improve contrast at lower energies^30^ and separate materials of different densities at higher energies while also reducing the deleterious effects of high density materials, i.e. calcium blooming and metal artifacts^31–34^.

The first clinical PCCT combines dual source CT and photon-counting detectors. This study assessed the stability of spectral results of a clinical dual-source PCCT for different acquisition modes and cardiac phases using a cardiac motion phantom with elements simulating a coronary lumen with and without a calcified stenosis.

## Materials and Methods

### Phantom

To evaluate the effect of acquisition mode, cardiac phase, and heart rate on spectral results, a cardiac motion simulator (Sim4D-VL, Quality Assurance in Radiology and Medicine, Möhrendorf, Germany) that replicated coronary artery movement was utilized (Figure 1)^35^. A set of rods mimicking coronary stenoses of different materials and severity was submerged in water and surrounded with a cardiac phantom (Cardio QRM, Quality Assurance in Radiology and Medicine, Möhrendorf, Germany) and extension ring (Figure 1B). Specifically, analysis was performed for the stenosis with a 50% eccentric calcium plaque measuring 1.5 mm diameter and comprising 373 mg/cm^3^ hydroxyapatite (Figure 1C).

**Figure 1.**
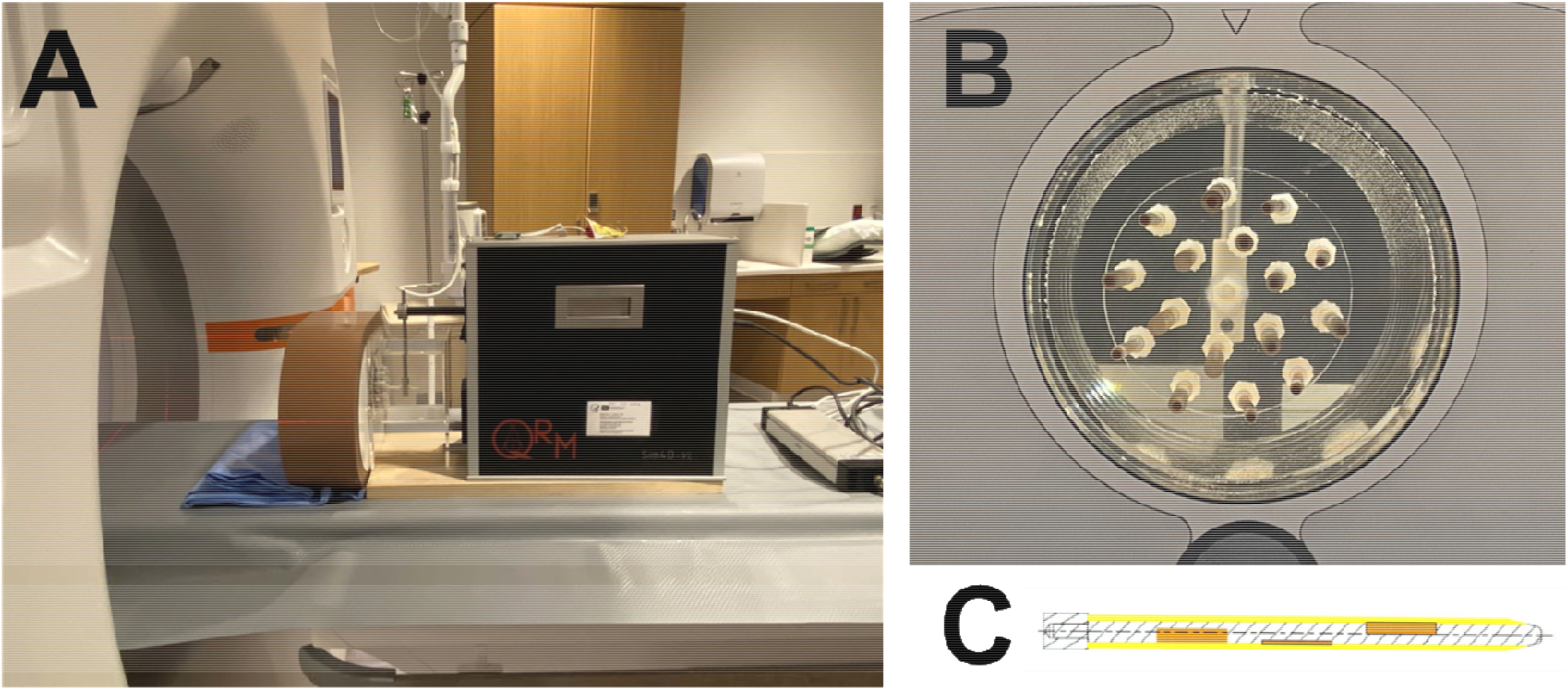
Experimental set up. A cardiac motion phantom was scanned with a dual-source photon-counting CT (A). The phantom included rods mimicking coronary stenoses of different materials (B) and extents (C).

### Image acquisition

The phantom was scanned with a first-generation dual-source PCCT (NAEOTOM Alpha, Siemens Healthineers, Erlangen, Germany) in the three available ECG-synchronized modes: dual-source low-pitch retrospective helical (spiral), dual-source prospective step-and-shoot (sequence), and dual-source high pitch prospective helical (flash) at different heart rates (0, 60, 70, 80, and 100 beats per minute). Scans were performed at 120 kVp at a fixed effective exposure of 30 mAs to match noise between scan modes (∼22 HU on VMI 70 keV). This exposure corresponded to average volumetric CT dose index (CTDI_vol_) of 20, 13, and 2 mGy for spiral, sequence, and flash, respectively. While both spiral and sequence scans allowed for reconstruction at systole (35%) and diastole (70%) with a single scan because scans encompassed systole to diastole (35 – 70%), separate flash scans were required for cardiac phases of 35% and 70%. To ensure the stenosis was imaged at 35% and 70% with flash scans, the location of the center of the stenosis was determined relative to the scan range, and the cardiac phase range at each heart rate was adjusted to ensure the center of the stenosis was scanned at the appropriate cardiac phase (35% or 70%). Subsequently, at each acquisition mode, cardiac phase, and heart rate combination, VMI 50, 70, and 150 keV as well as iodine density maps were reconstructed with a slice thickness of 0.4 mm at 0.4 mm intervals, field of view of 200 mm, reconstruction kernel of Bv36, matrix size of 512 × 512, and quantitative iterative reconstruction (QIR) level of 2. These parameters matched clinical protocols used at our institution. Other relevant scan and reconstruction parameters are included in Table 1.

**Table 1.**
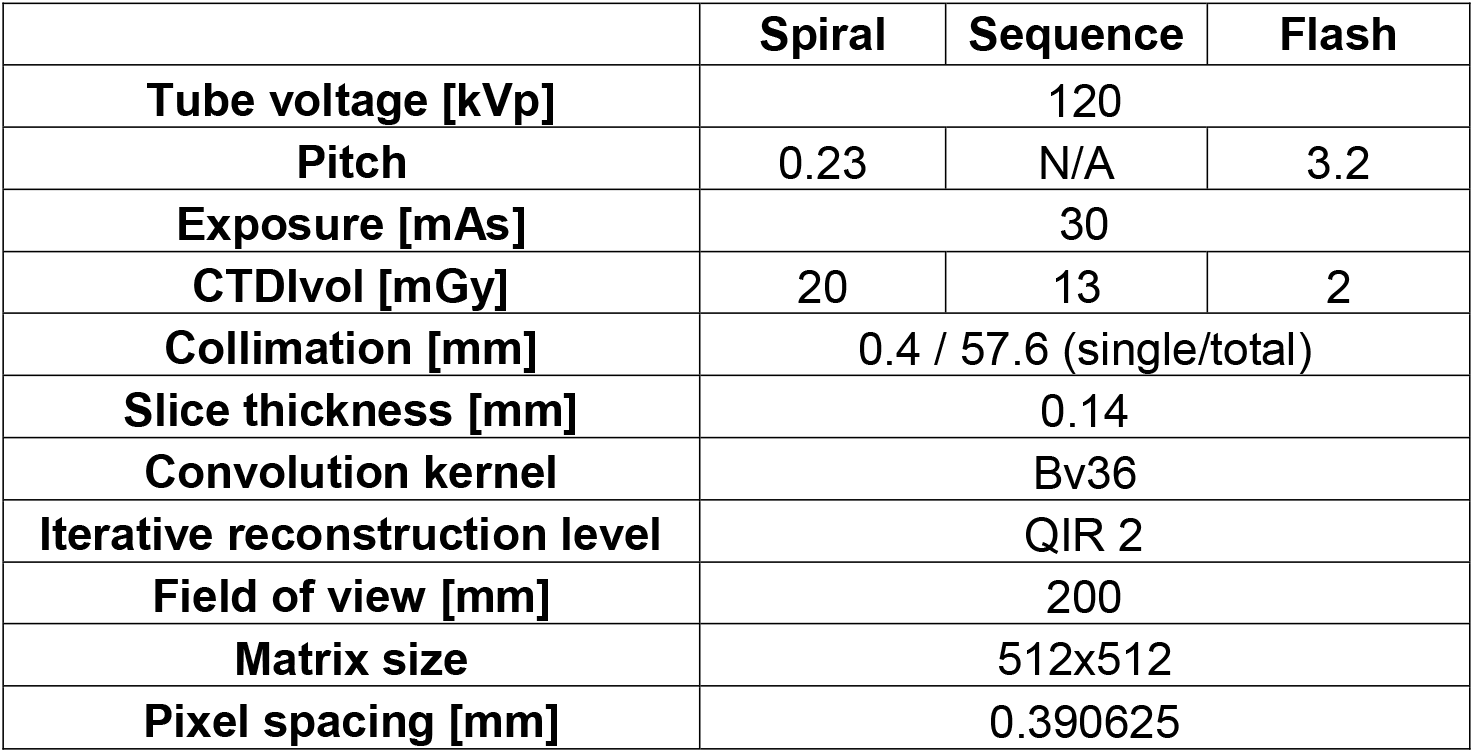
Acquisition and reconstruction parameters for different acquisition modes.

### Image analysis

To characterize the reliability of imaging of the stenosed and non-stenosed regions across acquisition mode, cardiac phase, and heart rate, quantitative metrics were investigated to describe size and similarity of the stenosed region as well as shape of the non-stenosed region. First, the center of the stenosis range was automatically determined. Because the lumen and calcified stenosis were similar on all spectral results except VMI 150 keV, adaptive thresholding was applied to the imaging volume of VMI 150 keV with adaptiveThreshold from the OpenCV package^36^. The range of slices with the stenosis was determined and the center calculated. Then line profiles at the center of the stenosis were evaluated to establish the similarity in width at different acquisition modes, cardiac phases, and heart rates. From the axial slices, line profiles were extracted, and the main peak corresponding to the stenosis was isolated by a threshold determined by the mean + standard deviation of the first 5 and last 5 values of the line profile. Once the peak was isolated, the full width half max (FWHM) was calculated for each set of parameters and spectral results. FWHM was reported as mean and standard deviation over 5 slices. Values were plotted for each spectral result individually, and average differences relative to the static FWHM were calculated.

For determining the similarity to static scans, five central axial slices of the stenosis were subjected to adaptive thresholding at each acquisition mode, heart rate, and phase for each spectral result to generate a mask. The mask for the static case (0 bpm) was then utilized to calculate the Dice similarity coefficient (DSC) for scans at the corresponding acquisition mode but different heart rate and cardiac phase, including another static scan (Figure 2). For each set of parameters, DSC was reported as mean and standard deviation across the five slices for each spectral result and visualized in a scatter plot. Standard deviation across all parameters for each spectral result as well as standard deviation across heart rate for combinations of spectral result, cardiac phase, and acquisition mode were also calculated.

**Figure 2.**
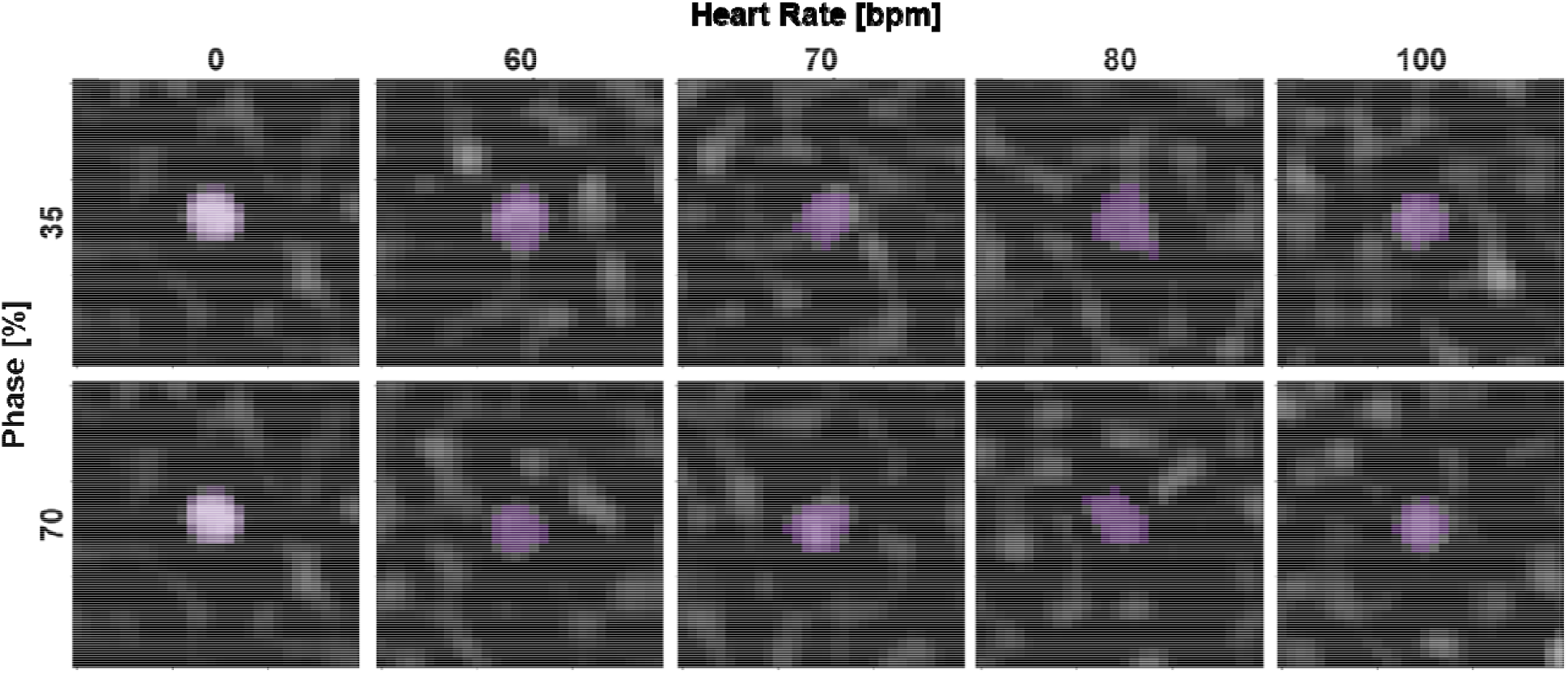
Adaptive thresholding of stenosis on VMI 150 keV images at varying cardiac phases and heart rates. Stenoses at different phases and heart rates were isolated with adaptive thresholding (purple) and demonstrated similar shape and size in comparison to the static scan (0 bpm).

Similarly, the non-stenosed regions were assessed for consistency between static and dynamic scans, by comparing eccentricity of the rod from the static circle. Eccentricity also served as an indicator of motion blurring. Eccentricity was only evaluated on VMI 50 and 70 keV and iodine density maps but not for VMI 150 keV. As iodine contrast reduces at higher VMI energies, the non-stenosed region on VMI 150 keV was not distinguishable from the background (water). Therefore, adaptive thresholding was applied to five axial slices at VMI 50 and 70 keV and iodine density maps. The contour of the generated mask was then fitted to an ellipse to determine the major and minor axes (Figure 3). Eccentricity was then calculated as:

**Figure 3.**
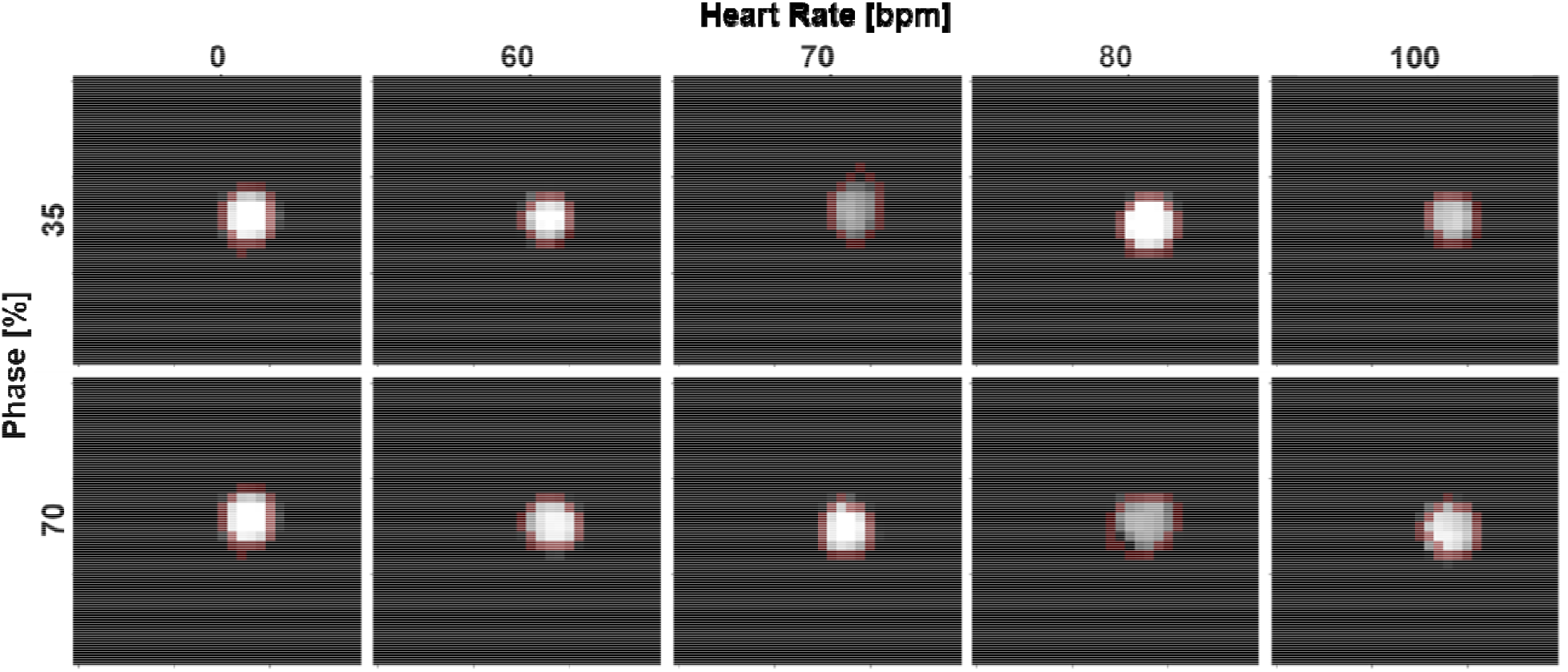
Shape evaluation of shape for non-stenosed region of the rod. Ellipse (red) were fitted to rods isolated from adaptive thresholding. At different phases and heart rates, the shape of the rod remained the same with some distortion at higher heart rates.

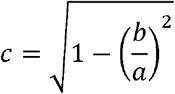

where c is eccentricity, b is radius of the minor axis, and a is the radius of the major axis. An eccentricity of 1 denoted a line, while an eccentricity of 0 denoted a circle. It was represented in a scatter plot with mean and standard deviation across 5 slices for each spectral result, acquisition mode, cardiac phase, and heart rate combination. The range of dynamic values and average difference from eccentricity measured in the static scan were evaluated. All image processing and analysis were performed in Python (Python Software Foundation).

### Statistical analysis

For all three metrics, Shapiro-Wilk tests were used to test for normality. Then, a three-way analysis of variance (ANOVA) was performed to assess the effect of acquisition mode, cardiac phase, and heart rate. Static data (0 bpm) was omitted as no motion was present, and thus did not provide an adequate comparison. A p value less than 0.05 was considered significant. Because three-way ANOVAs revealed a three-way interaction effect, additional simple two-way ANOVAs were utilized to determine the effect of acquisition mode and heart rate for each cardiac phase. A p-value of 0.025 was the metric of significance after Bonferroni correction. Simple main effects via one-way ANOVA were also performed to separate two-way interaction effects by evaluating the effect of heart rate for combinations of acquisition mode and cardiac phase. A p-value less than 0.0167, which was adjusted for Bonferroni correction, was considered significant and reported. All statistical analysis was performed in R (R Core Team) and RStudio (RStudio Team).

## Results

FWHM at different heart rates and acquisition modes demonstrated consistency across different spectral results and cardiac phases (Figure 4). This consistency is demonstrated by the small average differences from the static FWHM of -0.20, -0.28, and -0.15 mm for VMI 150 keV of flash, sequence, and spiral scans, respectively, and reflected that the size of the stenosis was preserved at different parameters. The size of the rod did not vary much, with average differences ranging from -0.02 to 0.32 mm and larger differences seen on iodine density maps. Statistical testing corroborated that there was no effect of heart rate and acquisition mode at diastole (p-values less than 0.001). In particular, the effect of heart rate was not significant for spiral 35%, flash 70%, sequence 70%, and spiral 70% for each of the spectral results. While other parameters demonstrated significant effect with heart rate, differences in FWHM were small, corresponding to a maximal 13% difference from the static measurements. Despite the consistency for each spectral result, the overall magnitude of the FWHM was smaller for VMI 150 keV (1.8 mm) in comparison to other spectral results (2.4, 2.3, and 2.6 mm for VMI 50 keV, VMI 70 keV, and iodine density maps, respectively). The difference corresponded to the reduced contrast of the iodine-containing lumen in VMI 150 keV, isolating the stenosis compared to other spectral results.

**Figure 4.**
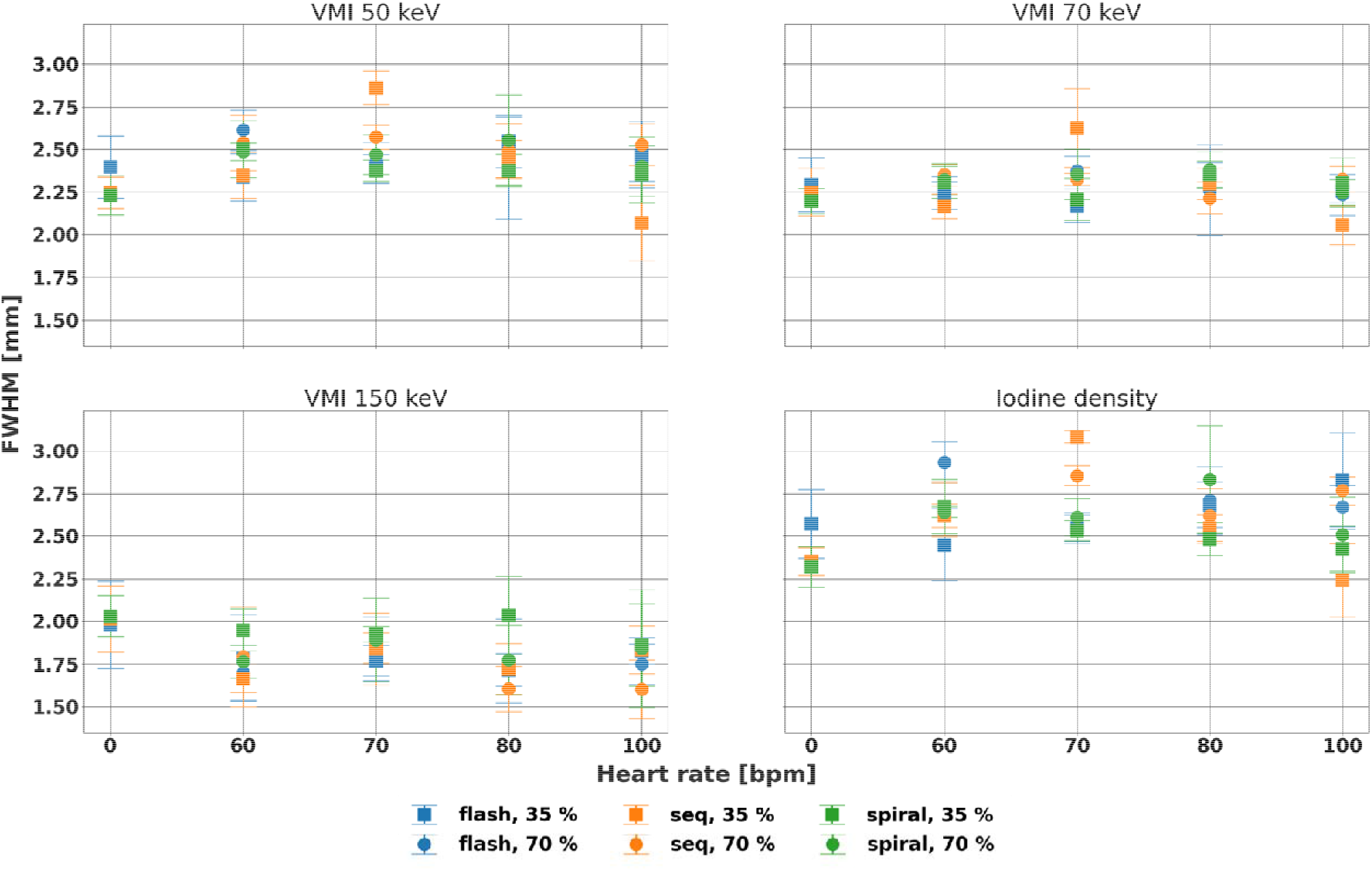
Consistency of full width half max (FWHM) for line profile of stenosed region with various parameters. FWHM varied with heart rate at systole for VMI 50, 70 keV, and iodine density maps but not vary with heart rate or acquisition mode at diastole for all four spectral results. Differences from the static measurements were small at a maximum of 13%.

DSC proved mostly similar for each spectral result and acquisition mode with respect to heart rate but showed larger differences at systole and with flash scans (Figure 5). Across acquisition mode, cardiac phase, and heart rate, standard deviation measured 0.08, 0.09, 0.11, and 0.08 for VMI 50, 70, and 150 keV, and iodine density maps, respectively. Specifically, for VMI 150 keV, where the stenosed region was best separated from the lumen, DSC maintained its value across dynamic scans with standard deviations spanning 0.14, 0.15, 0.11, 0.09, 0.10, and 0.08 for flash 35%, flash 70%, sequence 35%, sequence 70%, spiral 35%, and spiral 70%, respectively. The minor effect of acquisition mode was also exhibited for VMI 50 and 70 keV, and iodine density maps. Notably, some of the variation for each acquisition mode and cardiac phase combination can be attributed to specifically a heart rate of 80 bpm rather than higher heart rates, which can be observed both qualitatively and quantitatively. This is a well known effect given the reduced cardiac motion at heart rates greater than 80 bpm. While DSC significantly differed among acquisition modes for different cardiac phases and spectral results (p-values < 0.01), the effect of heart rate was not significant for spiral 35%, sequence 70%, and spiral 70% for any evaluated spectral results, demonstrating the stability at diastole and with spiral mode, with larger differences in systole and with flash scans.

**Figure 5.**
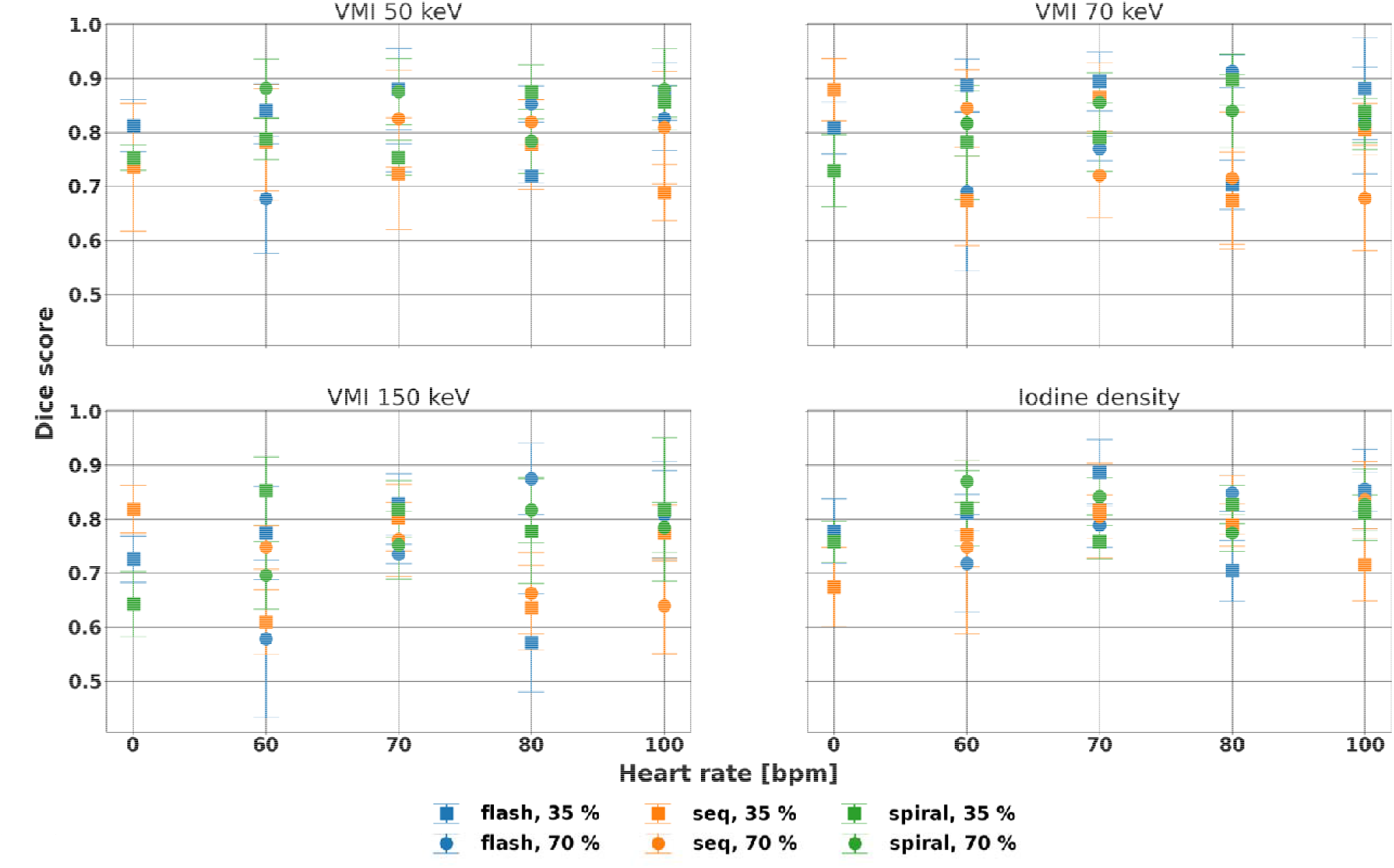
Stability of Dice score of the stenosed region for different spectral results. Dice score demonstrated different primarily associated with heart rate at systole. Clear variation in Dice score at 80 bpm was present, particularly for VMI 70 and 150 keV.

Similarly, shape of the rod in non-stenosed regions as represented by eccentricity remained stable at different heart rates and acquisition modes at varying cardiac phase and spectral results (Figure 6). Across parameters, eccentricity differences from the static scan averaged 0.012, -0.021, and 0.103 for VMI 50 and 70 keV and iodine density maps, respectively. At VMI 70 keV, the range of differences relative to the static value at diastole were 0.121, 0.274, and 0.168 for flash, sequence, and spiral, indicating that similarity to the static scan depends on acquisition mode. Furthermore, at systole, ranges were larger at 0.157, 0.224, and 0.183 for flash, sequence, and spiral, respectively. Even though eccentricity at systole and diastole were dependent on heart rate (3/3 and 2/3 spectral results, respectively), it was not dependent on acquisition mode (1/3 and 0/3 spectral results, respectively). In particular, heart rate had a significant effect on eccentricity for sequence 35% at VMI 50 keV, spiral 35% at VMI 50 keV, and sequence 70% at VMI 70 and iodine density maps (p-values < 0.01). Even so, differences in eccentricity remained small compared to eccentricity with static scans.

**Figure 6.**
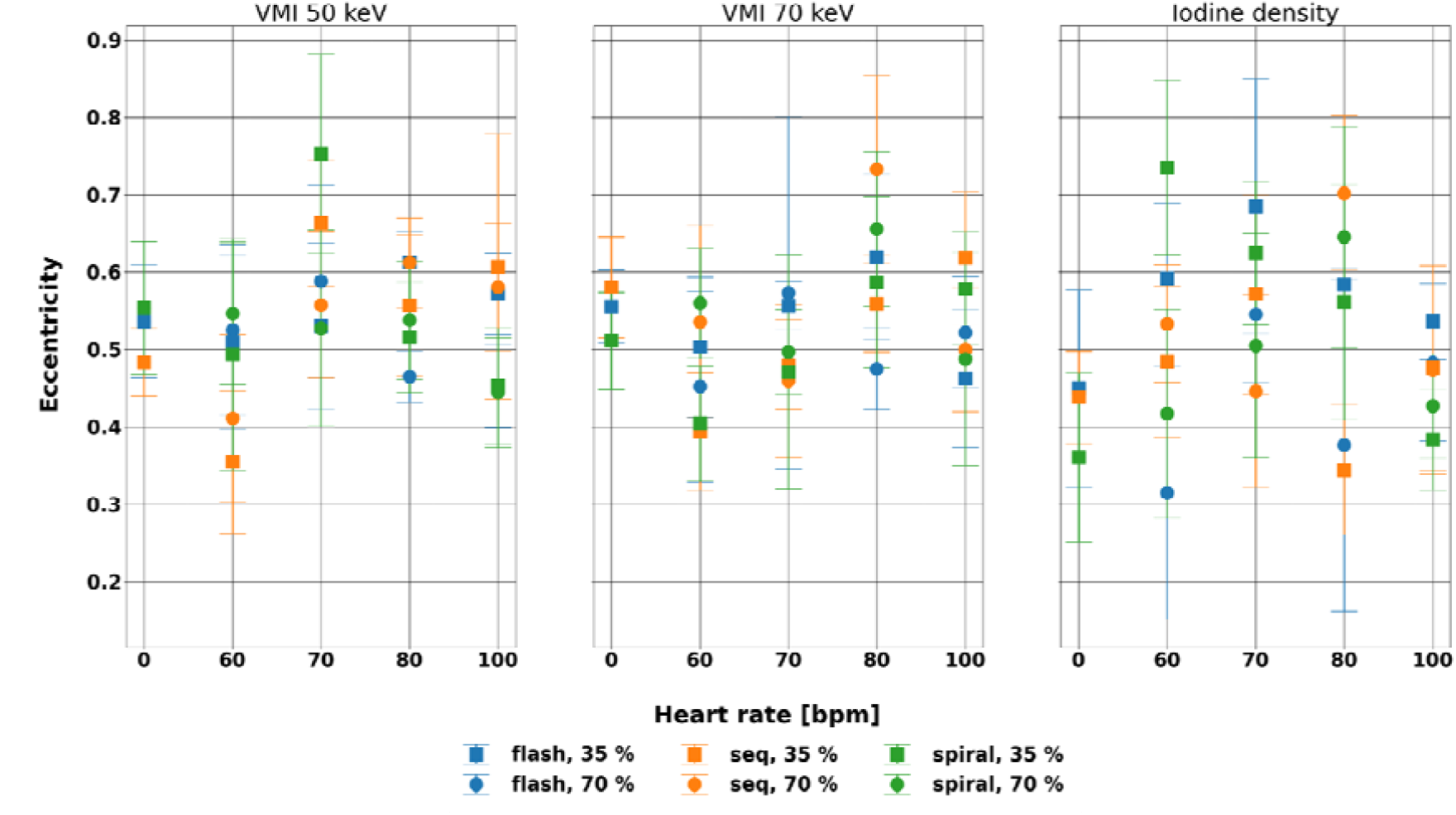
Eccentricity of the non-stenosed region of the rod at different spectral results for different acquisition mode, phase, and heart rate. Eccentricity was not affected by acquisition mode at both systole and diastole while was affected by heart rate, especially for sequence 35% at VMI 50 keV, spiral 35% at VMI 50 keV, and sequence 70% at VMI 70 keV and iodine density maps.

## Discussion

Spectral results exhibited stability with heart rate and acquisition modes at different cardiac phases, with more stability in diastole. The PCCT scanner improves upon existing dual-source dual energy CT with the addition of spectral results while maintaining the image quality available with dual-source CT. This systematic evaluation of the effect of heart rate and acquisition mode on spectral results provides confidence in the reliability of spectral in cardiac CT exams.

With the addition of energy discriminating detectors to dual source CT in the first clinical PCCT, its performance for cardiac CT with respect to heart rate and phase was similar to previous generations of dual source CTs. In spiral mode with second generation dual source CT, the optimal cardiac phase for reconstruction peaked at 30% and 80% between 50 and 100 bpm with measurement overestimation for diameter ranging from 0.1 to 0.4 mm depending on the stenosis size, comparable to FWHM variation across all phase and acquisition mode combinations with PCCT (0.003 to 0.37 mm)^37^. In particular, for heart rates less than 70 bpm, reconstructions at 70% performed the best with the least motion artifact compared to other evaluated phases^15^, while at higher heart rates, there is competing data in regards to the optimal phase^15,37^. While we did not directly evaluate motion artifacts and investigated various spectral results rather than conventional images in this study, results at diastole demonstrated consistent size and shape across heart rate and additionally at systole with spiral mode with the exception of eccentricity at VMI 50 keV. For flash mode, diagnostic image quality was previously reported to be available up to 75 bpm with more artifacts at higher heart rates with lower dose^38^. Likewise, quantitative evaluation via FWHM, DSC, and eccentricity exhibited variation with heart rate with flash, both at diastole and systole, with deviations at higher heart rates. The performance with respect to heart rate highlights that the temporal resolution benefits of dual source PCCT were comparable but with the additional availability of spectral results.

While spectral results have been available with other CT technology, the combination of dual source and photon counting CT enables spectral results at higher temporal and spatial resolution in addition to radiation dose reduction. The stability of VMI 50, 70, and 150 keV, and iodine density maps, it can be extended to other spectral results, provides confidence in both quantification and visualization of coronary arteries and other structures. In particular, these spectral results are important for coronary CT angiography, which has increasingly played a role in diagnostic cardiac imaging over invasive angiography^8–10,39–41^. Both virtual monoenergetic images and effective atomic number maps can characterize the composition of the plaque, i.e. calcification, fibro-fat, and fat, and help better estimate the extent of obstruction for calcified plaques that previously were overestimated as a result of calcium blooming^31,42,43^. Reduction of calcium blooming artifacts may also permit the evaluation of myocardial ischemia that may result from coronary obstruction, which previously was limited by such artifacts and motion artifacts^33,34^. Generally, accurate identification and categorization of the plaque may guide more refined risk stratification and improve clinical care. In addition to traditional evaluation of coronary artery stenosis with coronary CT angiography, it was feasible to perform calcium scoring with virtual non contrast images from coronary CT angiography, eliminating the need for a dedicated non-contrast scan^28,29^. Late iodine enhancement cardiac CT in both dual-layer CT and PCCT illustrated good agreement with magnetic resonance imaging in terms of both visualization with VMI and iodine density maps and quantification of the extracellular volume important for evaluating myocardial fibrosis, edema, and viability in cardiomyopathy^23–25^. Virtual non contrast images, on the other hand, have exhibited improved preservation of stent and calcified structures compared to conventional CT for the surveillance post endovascular aneurysm repair^44^. With the consistency of the spectral results of dual source PCCT, these applications can be investigated and implemented to improve cardiovascular diagnostics.

This study was limited in a few ways. First, in comparison to other studies, the analyzed stenosed region included a calcified stenosis similar in attenuation at low keVs to the lumen and, thus, low in contrast. The lumen and stenosis were best separated on VMI 150 keV as a result of the reduction in the contrast of the lumen in comparison to the stenosis. This provided a more challenging task for evaluating image quality and structural preservation and prevented evaluation of non-stenosed regions on VMI 150 keV. However, it demonstrated the advantages of spectral results, which highlighted the stenosis. Other studies have evaluated stenoses at standard 200, 400, and 800 mg/mL of hydroxyapatite either isolated or at lower background attenuation (200 HU), which provide more contrast^2,37^. Second, while the motion of the phantom was modeled off Achenbach et al., the phantom did not properly replicate cardiac motion at high heart rates (> 80 bpm) where cardiac motion reduces rather than increases. Despite this, the phantom provided flexibility with inserts comprising of stenoses of different materials and extents. A single clinically-relevant stenosis was only analyzed in this study to demonstrate the stability of spectral results. Third, only one phantom size was investigated in this study. Different patient habitus may affect quantification as has been demonstrated with static phantoms, but it is unknown how it may affect dynamic scans.

In conclusion, heart rate and acquisition mode did not affect the size of the stenosis, similarity to the static scan, and shape of the non-stenosed region at diastole for different spectral results. With improved material characterization and quantification, these spectral results may ultimately contribute to the advancement of cardiovascular diagnostics via enhanced cardiac CT imaging protocols.

## Data Availability

All data produced in the study are available upon reasonable request to the authors.

## Acknowledgements

We acknowledge support through the National Institutes of Health (R01EB030494) and Siemens Healthineers.

